# Corticospinal and Reticulospinal-Associated Measures in Chronic Ischemic Stroke: Associations with Upper Limb Function

**DOI:** 10.64898/2026.10.26.26365898

**Authors:** Asit Baran Bayen, Subhajit Sarkar, Sucheta Saha, Uttara Bakshi, Chiranjib Das, A. Shobhana, Anirban Ghosal, Soumava Mukherjee, Siddharth Shankar Anand, Arijit Pal, Apratim Chatterjee, Suparno Gangopadhyay, Sanjit Dey, Purba Basu, Boubker Zaaimi, Mark R. Baker, Stuart N. Baker, Hrishikesh Kumar, Supriyo Choudhury

## Abstract

**Background:** Corticospinal tract (CST) damage is the main cause of post-stroke motor impairment. Whilst the output of the reticulospinal tract (RST) is reportedly upregulated, its relation to CST damage post-stroke is unclear. We assessed CST integrity using transcranial magnetic stimulation and RST activity using the StartReact paradigm in chronic ischemic stroke and examined their inter-relationship and associations with upper-limb impairment.

**Methods:** We included 80 patients with chronic ischemic stroke and 15 age-matched healthy controls (HCs). CST function was quantified using the resting motor threshold asymmetry index (RMT-AI); RST function was measured by the StartReact effect (visual-auditory minus visual-startle reaction time) during isometric wrist flexion. Impairment was assessed with the Fugl-Meyer Assessment-Upper Extremity (FMA-UE) and affected-side grip strength, with FDR-corrected correlations.

**Results:** Higher RMT-AI was associated with lower FMA-UE (r −0.46, p 0.0038) and weaker grip (ρ −0.43, p 0.0065). A larger StartReact effect showed the same pattern (ρ −0.35, p 0.0095, FMA-UE; ρ −0.41, p 0.00195, grip). All correlations remained significant after FDR correction. RMT-AI and the StartReact effect were uncorrelated (r 0.05, p 0.792). The StartReact effect did not differ between sides (p 0.631) or between patients and HCs (p 0.342) but was larger in patients with FMA-UE ≤ median than in HCs (p 0.025), without a side difference (p 0.143).

**Conclusions:** Greater impairment was associated with greater CST damage and RST activity, possibly reflecting cortico-reticular fibre damage. CST and RST measures were unrelated, suggesting cortical input to the reticular formation beyond M1. Severely impaired patients showed greater bilateral reticulospinal involvement.

## Introduction

Stroke is a leading cause of long-term disability, and difficulty using the arm and hand is one of its most common and troublesome consequences. Even after rehabilitation, many people are left with weaknesses, poorly controlled movements, and difficulty moving one joint independently months or years after their stroke (1). The degree of recovery also varies widely, even between people with similar types and severity of stroke. This suggests that recovery depends not only on how much of the brain was damaged, but also on which motor pathways survive and how much each of them contributes to movement afterwards (1, 2).

In humans, the main pathway for voluntary movement of the arm and hand is the corticospinal tract (CST), which carries signals from the brain’s motor area to the spinal cord. It is especially important for fine, independent finger movements (2). The extent to which the CST is preserved has been linked to upper limb recovery after stroke, with greater damage to this pathway generally associated with greater motor impairment (3-5). However, CST measures alone do not fully explain recovery. Some people with no measurable response from this pathway still regain some strength and gross arm movement (6, 7), suggesting that other pathways may also contribute.

One of the likely candidates is the reticulospinal tract (RST) which reorganizes significantly after stroke. It starts in the brainstem, receives input from both sides of the brain, and sends signals to both sides of the spinal cord (8, 9). The RST was long thought to control mainly posture and the shoulder and upper arm. However, animal studies show that it also reaches the spinal neurons controlling the forearm and hand (10, 11), and that these connections grow stronger during recovery after CST damage (12). In humans, the two pathways may play different roles, with studies suggesting that the CST is involved particularly in precise finger control and the RST in strength and whole-hand grasp (13, 14). Relying more on the RST after stroke may therefore restore some gross movement. However, because it cannot control individual muscles necessary for fine motor control, some have suggested that it may also contribute to the abnormal movement patterns that limit everyday arm use (15-17), although animal studies contradict this idea (18). Whether greater involvement of the RST is helpful, harmful, or both remains an area of uncertainty (19, 20).

The RST is difficult to study in people because it cannot be measured directly. One practical approach makes use of the startle response. When a person is ready to make a quick movement in response to a signal, a sudden loud sound makes the movement start much sooner (21, 22). This speeding-up is thought to rely mainly on the RST (23), so its size gives an indirect estimate of how much the RST contributes to movement. The effect is preserved after stroke, and a startling sound can trigger fast movements of the affected arm and hand (24-26). Our group has previously shown that in people with chronic stroke, those with a larger RST contribution to movement tend to have poorer upper limb function (27) and a reduced ability to straighten the elbow when reaching (28). However, the CST contribution was not quantified in the same cohort.

Studying both pathways in the same group has already proved useful in other neurological conditions. After spinal cord injury, the RST appears to contribute more to whole-hand grasp than to fine pinch (29). Testing both pathways in these patients has shown that the CST and RST play different roles in controlling arm muscles (30). Greater RST activity has also been reported in hereditary spastic paraplegia, where the CST gradually degenerates (31). Together, these studies show that looking at both pathways at once can reveal their mutual relationship in relation to the residual motor impairment.

In stroke, only a few studies have examined both pathways together, and their findings, in relatively small cohorts of patients, do not fully agree. One study found that a stronger CST was linked to better muscle control, whereas a stronger RST was linked to greater strength (20). Another found that increased RST activity was seen mainly in people with severe CST damage (7). In contrast, a study in the first months after stroke reported that upper limb impairment was related to CST rather than RST function (32).

Several important questions therefore remain. Most studies have assessed either the CST or the RST, but not in the same patients. CST function has often been recorded simply as present or absent, rather than graded by comparing the damaged and healthy sides of the brain. Previous studies of the RST in stroke have also been relatively small and have mostly relied on overall measures of upper limb function. They have not examined specific aspects of impairment, such as hand function, grip strength, or the ability to move outside abnormal movement patterns. Most importantly, it is not known how the two pathways compare in their relationship with upper-limb impairment when both are measured in the same patients.

Therefore, we examined whether the integrity of each descending pathway correlated with overall upper-limb impairment and with grip strength in the affected hand. We also assessed how corticospinal and reticulospinal function correlate with each other within the same patients and compared reticulospinal function in patients with that of age-matched healthy controls.

## Methods

The study was conducted in the movement disorders laboratory of a tertiary care referral center in Eastern India.

### Consent to Participate and Publish

The study was approved by the Institutional Ethics Committee of Institute of Neurosciences Kolkata (protocol no. INK/ISH/TaVNS/(V-STIM)/2023/Ver.1.1, approved on 27 April 2023). Written informed consent was taken from all participants. The research was performed in accordance with the relevant guidelines and regulations.

### Inclusion and Exclusion Criteria for participants

Participants were eligible for inclusion if they were consenting male or female patients with a history of supratentorial ischemic stroke occurring at least six months but not more than five years prior to enrollment, and aged 30–70 years. Participants were required to have clinical history and neuroimaging evidence of ischemic stroke with residual post-stroke upper-limb motor impairment. Participants were also required to be on a stable treatment regimen during the study period.

Participants were excluded if they were wheelchair-bound or bedridden, had a history of hemorrhagic stroke, significant sensory impairment, concomitant neurological diseases such as tumor, epilepsy, demyelinating disease, vascular Parkinsonism or vascular dementia, or a history of more than two strokes. Patients with strokes involving two different cerebral hemispheres, uncontrolled systemic disease, pregnancy or lactation, uncontrolled or severe depression with suicidal tendency, global aphasia, significant hearing or visual impairment were also excluded. In addition, patients with severe upper-limb spasticity (Modified Ashworth Scale >3) were excluded.

Fifteen age-matched healthy controls with no history of neurological or musculoskeletal disorders affecting the upper limbs were also recruited. Written informed consent was obtained from all controls.

### Clinical Assessments

Comprehensive neurological and clinical assessments were performed in all patients with chronic ischemic stroke. Upper-limb motor impairment was assessed using the Fugl-Meyer Assessment for the Upper Extremity (FMA-UE), administered by a trained researcher (ABB) under the supervision of an experienced neuroscientist within our laboratory. Cognitive function was assessed using the Montreal Cognitive Assessment (MoCA).

Power grip strength was assessed with a Jamar-style electronic dynamometer (G200, Biometrics Ltd, Newport, UK), as in our previous work (27). Participants were seated with the shoulder adducted, the elbow flexed at 90° and the forearm in a neutral position. Three trials were performed with an interval of approximately 30 s, and the mean of the three trials (in kg) was used. The same protocol was repeated for the unaffected and affected sides.

### Experimental Protocol

#### Transcranial Magnetic Stimulation (TMS)

Single-pulse transcranial magnetic stimulation (TMS) was used to evaluate corticospinal excitability, delivered through a figure-of-eight coil (70 mm wing diameter) connected to a Magstim BiStim^2^ stimulator (Magstim Company Ltd, Whitland, UK). Motor responses were recorded with surface electromyography (EMG, Digitimer D360) from the first dorsal interosseous (FDI) muscle of both the affected and the unaffected hand. The active electrode was placed over the belly of the FDI, the reference electrode over the adjacent metacarpophalangeal joint, and the ground electrode on the back of the wrist. Signals were amplified with a gain of 500 and a digital lowcut filter of 5 Hz to high cut filter of 2000 Hz were used. EMG was then digitized at 5 kHz and saved to disk for offline analysis using a 1401 interface and Spike2 software (Cambridge Electronic Design, Cambridge, UK).

During testing, participants sat comfortably with the tested forearm and hand supported and relaxed. Stimulation was applied over the FDI representation of the primary motor cortex (M1). The coil was held tangentially to the scalp over the cortical hand area, with the handle pointing backwards and laterally at approximately 45° to the mid-sagittal line, so that the induced current flowed in a posterior–anterior direction across the central sulcus. The coil position was then adjusted until the largest and most reproducible MEPs were obtained. This optimal site was determined individually for each participant and kept constant for the rest of the session. Each hemisphere was tested separately, for the affected and unaffected sides.

##### Resting motor threshold

The resting motor threshold (RMT) was obtained with single-pulse TMS while the FDI muscle was fully relaxed. RMT was defined as the minimum stimulator output that evoked an MEP greater than 50 μV in at least 5 of 10 consecutive trials.

##### Central motor conduction time (CMCT)

CMCT was assessed by recording motor evoked potentials (MEPs) from the first dorsal interosseous (FDI/1DI) muscle, separately for the affected and unaffected sides. After the resting motor threshold (RMT) was established, 10 single TMS pulses were delivered over M1 at 120% RMT, and cortical MEP amplitude was taken as the mean of the 10 responses. To obtain a response independent of cortical stimulation, cervical MEPs were also recorded: a figure-of-eight coil was placed over the C7 region, with the neck flexed forward for positioning, and 10 stimuli were delivered on each side. MEP onset latency was measured for each of the 10 cortical and 10 cervical responses, and the mean of the 10 values was used. Total motor conduction time (TMCT) was defined as the mean onset latency of the cortical MEPs, and peripheral motor conduction time (PMCT) as the mean onset latency of the cervical MEPs. CMCT was then calculated for each side as the total motor conduction time minus the peripheral motor conduction time (CMCT = TMCT − PMCT). Recording both cortical and cervical responses on each side allowed comparison of corticospinal output between the two limbs.

##### Neurophysiological asymmetry indices

For measures recorded on both sides, an asymmetry index (AI) was used to express the difference between the affected and unaffected sides. The AI was calculated as:

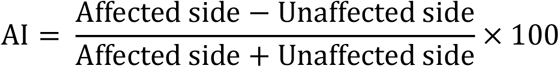

This formula was applied to obtain the RMT asymmetry index (RMT-AI) and the MEP magnitude asymmetry index (Magnitude-AI). A positive AI therefore indicates a higher value on the affected side, and a negative AI a higher value on the unaffected side (33).

### StartReact Experimental Protocol

The StartReact is a reaction time paradigm considered as an indirect measure of reticulospinal activity (23). We used the StartReact paradigm to measure how quickly participants responded to a visual cue paired with a low intensity sound, and how much this reaction time was shortened when the visual cue was paired with a startling sound. The procedure, including the apparatus, stimulus parameters and recording settings, followed the protocol previously established in our laboratory for studying reticulospinal contributions to upper-limb motor function in chronic stroke (27, 28).

Participants sat comfortably in a chair with the tested arm resting on the experimental apparatus. The forearm was supported and held steady, and the hand was placed in a semi-pronated position between two adjustable vertical plates. Participants were asked to react as fast as possible to a visual cue by making an isometric wrist-flexion movement. Response onset was identified from surface electromyography (EMG) recorded over the forearm flexor carpi radialis (FCR) muscle (Figure 1A).

**Figure 1.**
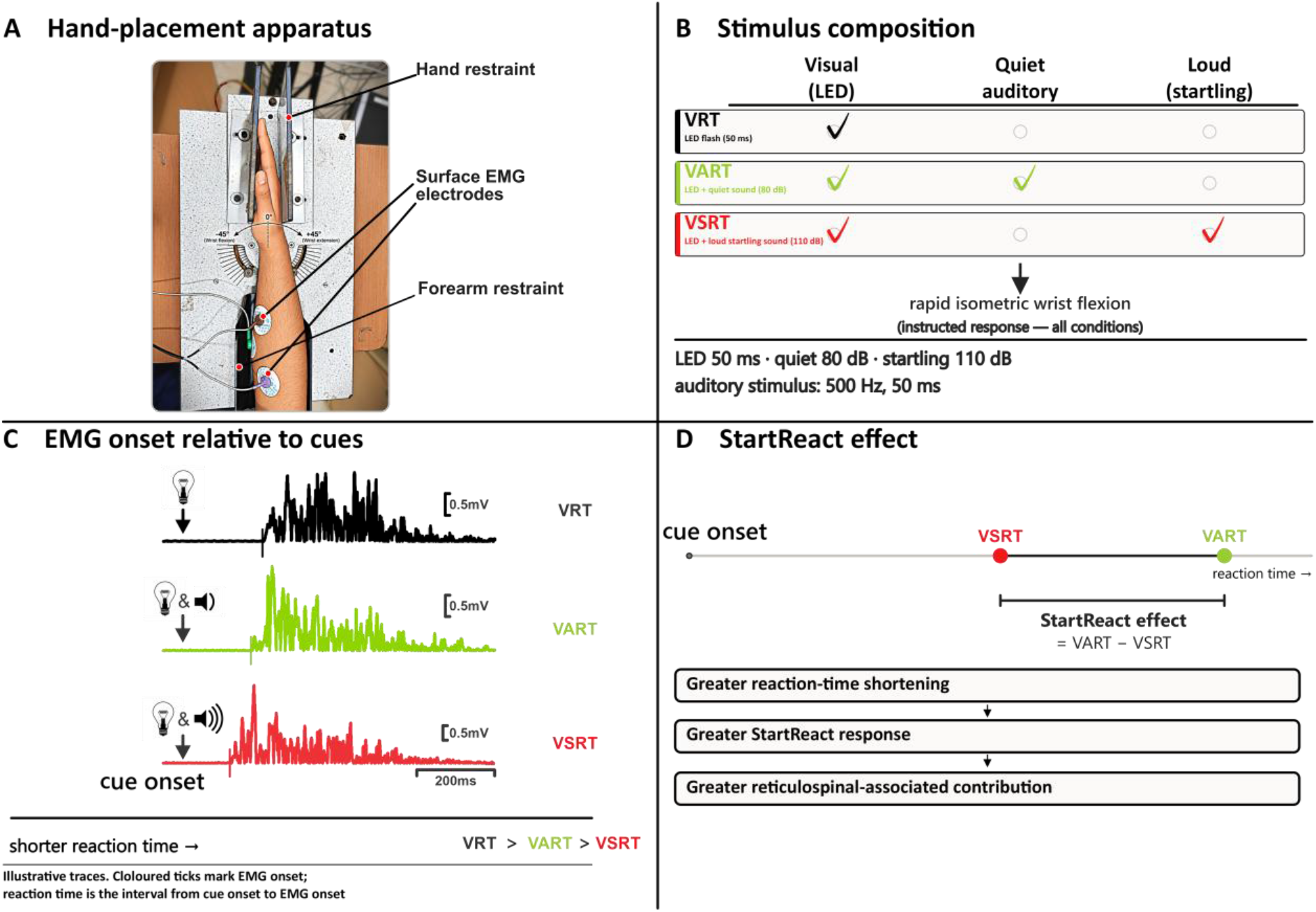
Schematic representation of the StartReact experimental paradigm and representative responses. **(A)** Experimental setup illustrating stabilization of the hand and forearm and placement of surface electromyographic (EMG) electrodes during the StartReact task. **(B)** Illustration of the three stimulus conditions: visual cue alone (VRT), visual cue accompanied by a non-startling auditory stimulus (VART), and visual cue paired with a startling auditory stimulus (VSRT). **(C)** Representative surface EMG responses recorded from the forearm flexor following VRT, VART, and VSRT stimulation. **(D)** Schematic depiction of the reduction in reaction time between VART and VSRT conditions, representing the StartReact effect, calculated as VART − VSRT; a greater reduction in reaction time reflects a larger StartReact response and is interpreted as greater reticulospinal-associated motor contribution.

The visual cue was a light-emitting diode (LED) that illuminated for 50 ms. Three conditions were delivered in random order: (i) the visual cue alone; (ii) the visual cue together with a low-intensity sound; and (iii) the visual cue together with a high-intensity, startling sound. The low-intensity sound was delivered at 80 dB and the startling sound at 110 dB, each lasting 50 ms with 500 Hz frequency (Figure 1B).

Before recording began, participants practiced the task by responding to the visual cue. They were then habituated to the startling sound. Neither the practice nor the habituation trials were analyzed. We then recorded 20 trials of each of the three conditions, presented in random order with an interval of 5 s between trials. Surface EMG were recorded from the forearm flexor muscles using a bipolar montage.

Reaction time on each trial was measured from the onset of the visual cue to the onset of the EMG response. EMG onset was determined relative to the pre-stimulus baseline. All automatically detected onsets were then checked manually so that any artefacts could be identified and corrected. The analysis was performed using a MATLAB script. Reaction times in the three conditions were termed: the visual reaction time (VRT) for the visual cue alone; the visual–auditory reaction time (VART) for the visual cue with the 80-dB sound; and the visual–startle reaction time (VSRT) for the visual cue with the 110-dB startling sound (Figure 1C). The StartReact effect was calculated for each participant as VART – VSRT (Figure 1D). A larger value therefore indicates that the startling sound shortened the reaction time more than the low-intensity sound did.

In HCs, the StartReact protocol was performed with both hands, and the right hand (dominant) was used for comparison with patients. To examine patients with more severe motor impairment, a subgroup was defined as those with a total FMA-UE motor score (sections A– D) at or below the cohort median.

### Statistical Analysis

Parametric or non-parametric tests were chosen for each variable according to its distribution and characteristics. Continuous variables are reported as mean ± standard deviation (SD), and categorical variables as counts and percentages. All tests were two-tailed, with statistical significance set at p < 0.05. Spearman rank correlation was used to examine how the StartReact effect (VART − VSRT) related to upper-limb motor impairment and to handgrip strength of the affected side. The outcomes examined were the total Fugl-Meyer Assessment–Upper Extremity (FMA-UE) motor score and handgrip strength.

The TMS-derived resting motor threshold asymmetry index (RMT-AI) was related to the StartReact measures using either Pearson or Spearman correlation, as appropriate. The same approach was used to examine the relationship of RMT-AI with upper-limb motor impairment and handgrip strength. Each correlation coefficient is reported together with its 95% confidence interval (CI). The StartReact effect was compared between the affected and unaffected sides using a paired t-test, after checking the normality of the paired differences with the Shapiro– Wilk test.

The affected-side StartReact effect of patients was compared with that of healthy controls (right hand) using the Mann–Whitney U test, both for all patients and for the subgroup with more severe upper-limb impairment. The 95% CI of the median difference between groups (patients − controls) was reported as the effect size. In an additional analysis, the affected and unaffected sides were compared within this subgroup using a paired t-test.

To account for the multiple correlations tested, p values were adjusted using the false discovery rate (FDR) method (Benjamini–Hochberg procedure), and significance after correction was set at q < 0.05. Where relevant, the results report the correlation coefficient (ρ for Spearman or r for Pearson), its 95% CI, and both the unadjusted and the FDR-adjusted p value (p-FDR).

Because not all measures were available for every participant, each analysis used all participants with valid data for that pair of variables, and the number of participants (N) is reported for each analysis.

## Results

### Participants

In this study, we recruited 80 chronic ischemic stroke patients and 15 age-matched healthy controls (HCs). Demographic details are given in Table 1.

**Table 1.**
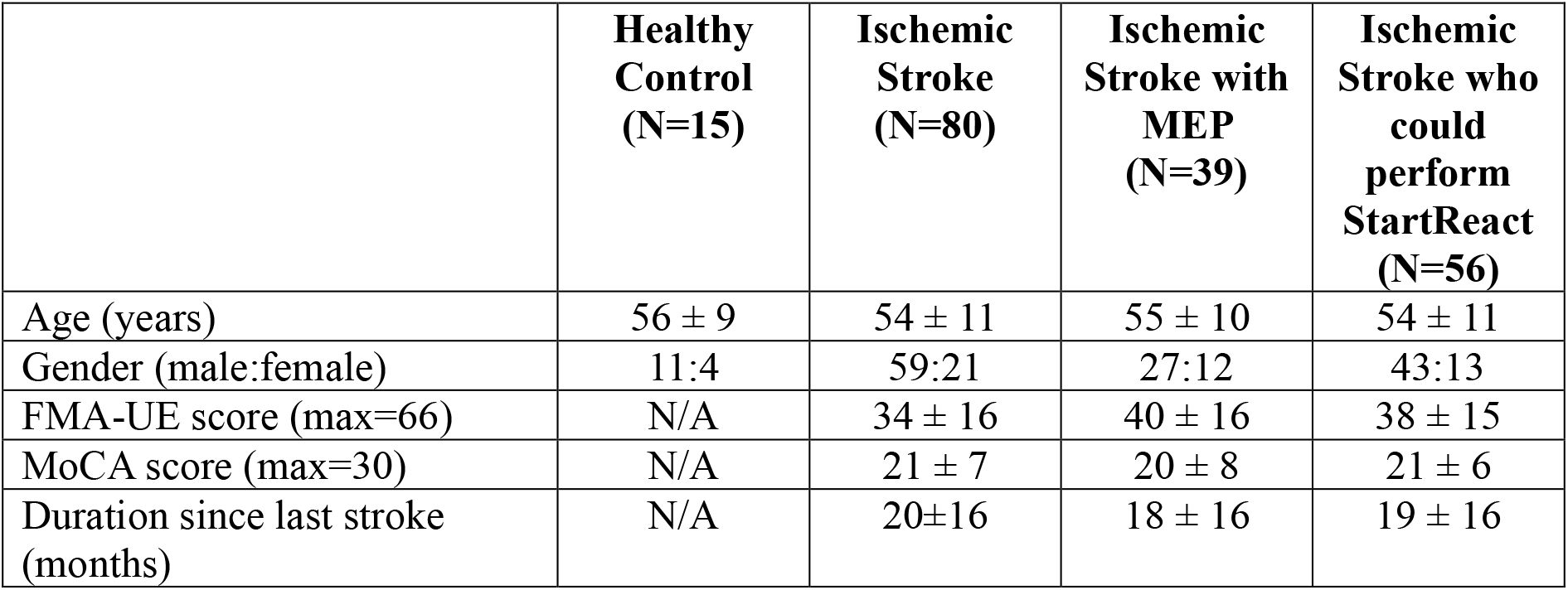
Demographic details, and clinical scores of participants. Values are given as Mean ± SD.

|  | **Healthy Control**<br>**(N=15)** | **Ischemic Stroke**<br>**(N=80)** | **Ischemic Stroke with MEP**<br>**(N=39)** | **Ischemic Stroke who could perform StartReact**<br>**(N=56)** |
| --- | --- | --- | --- | --- |
| Age (years) | 56 ± 9 | 54 ± 11 | 55 ± 10 | 54 ± 11 |
| Gender (male:female) | 11:4 | 59:21 | 27:12 | 43:13 |
| FMA-UE score (max=66) | N/A | 34 ± 16 | 40 ± 16 | 38 ± 15 |
| MoCA score (max=30) | N/A | 21 ± 7 | 20 ± 8 | 21 ± 6 |
| Duration since last stroke (months) | N/A | 20±16 | 18 ± 16 | 19 ± 16 |

### Association of corticospinal and reticulospinal-associated measures with upper-limb motor impairment

Higher RMT-AI was associated with lower total FMA-UE scores, indicating greater upper-limb impairment (Pearson’s r = −0.46, 95% CI −0.64 to −0.24; N = 38; p = 0.00385; p-FDR = 0.0103; Figure 2A), and with weaker affected-side grip strength (Spearman’s ρ = −0.43, 95% CI −0.68 to −0.11; N = 39; p = 0.00653; p-FDR = 0.0104; Figure 2B).

**Figure 2.**
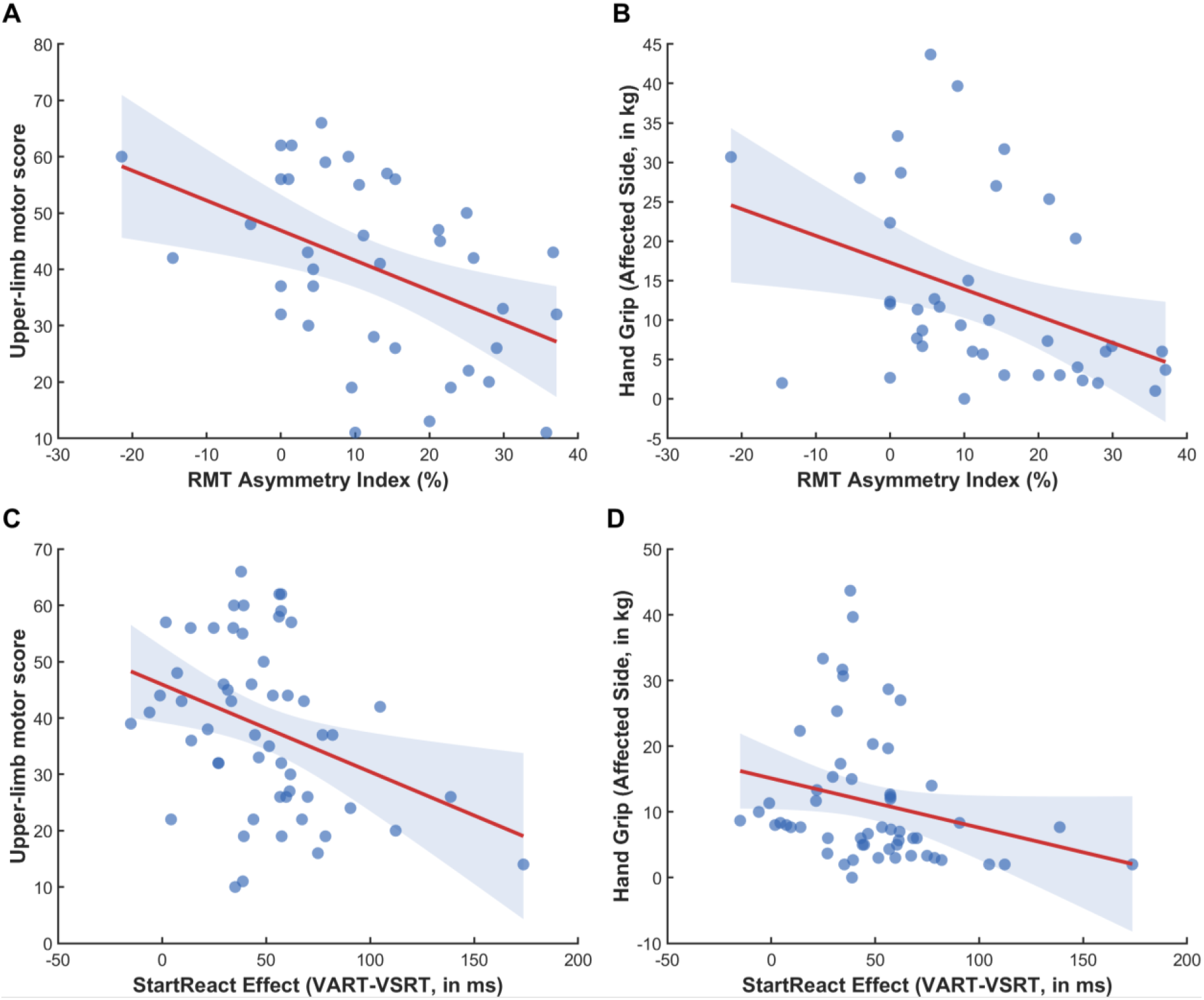
Association of corticospinal and reticulospinal-associated measures with upper-limb motor impairment and affected-side grip strength. Scatter plots show the relationship of the RMT asymmetry index with **(A)** the upper-limb motor impairment score and **(B)** affected-side handgrip strength, and of the StartReact effect (VART − VSRT) with **(C)** the upper-limb motor impairment score and **(D)** affected-side handgrip strength. Upper-limb motor impairment was assessed with the Fugl-Meyer Assessment–Upper Extremity (FMA-UE) motor score (maximum 66), in which higher scores indicate better motor function, i.e. less impairment. Regression lines with 95% confidence bands are shown.

Similarly, a larger StartReact effect was associated with lower total FMA-UE scores (Spearman’s ρ = −0.35, 95% CI −0.55 to −0.10; N = 55; p = 0.00946; p-FDR = 0.0252; Figure 2C) and with weaker affected-side grip strength (ρ = −0.41, 95% CI −0.59 to −0.18; N = 56; p = 0.00195; p-FDR = 0.00782; Figure 2D).

### Relationship between corticospinal and reticulospinal-associated measures, and comparison with healthy controls

RMT-AI was not associated with the StartReact effect in our cohort (Pearson’s r = 0.05, 95% CI −0.40 to 0.51; N = 27; p = 0.792; p-FDR = 0.792; Figure 3A).

**Figure 3.**
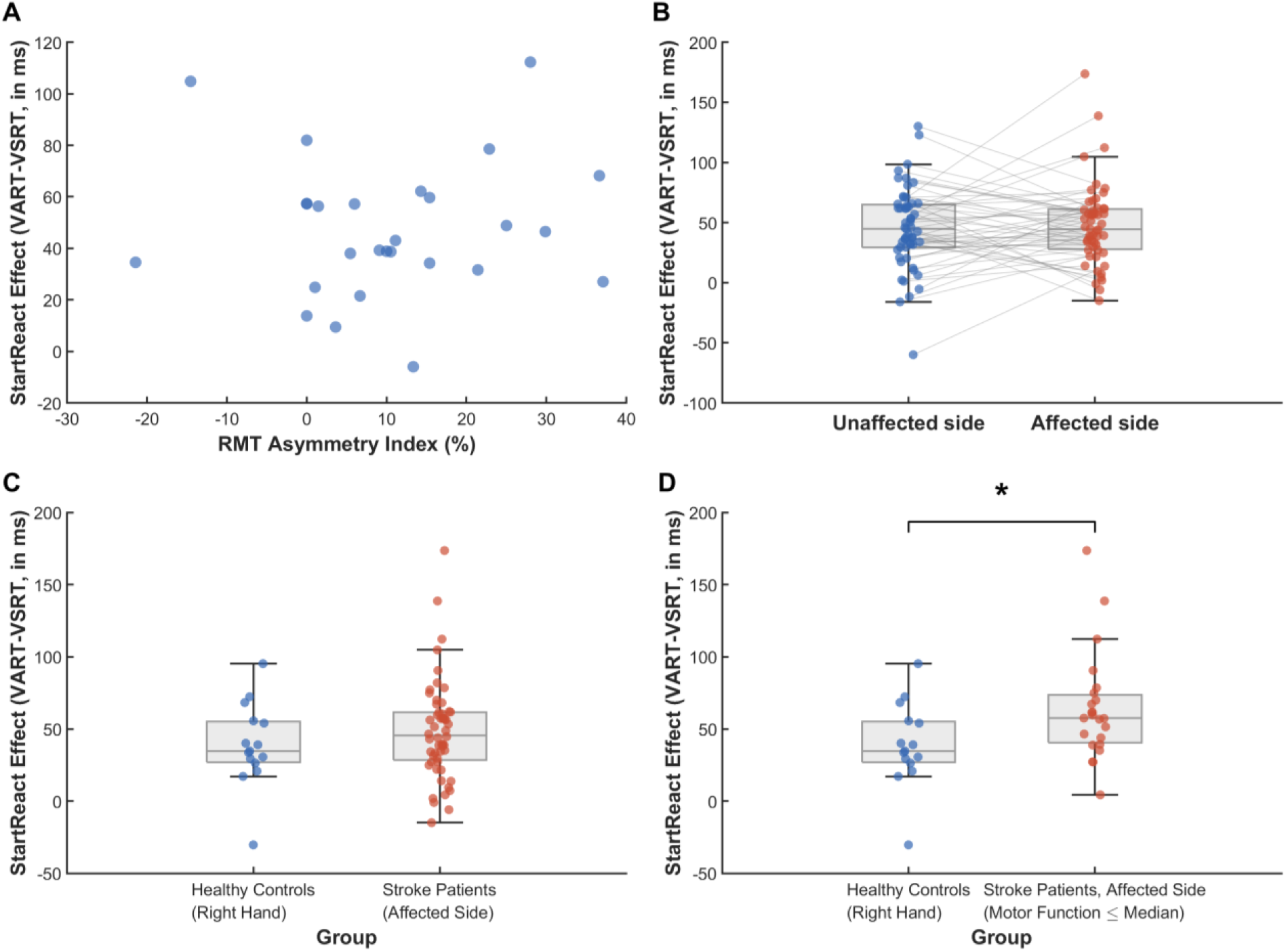
Relationship between corticospinal and reticulospinal-associated measures, and comparison with healthy controls. **(A)** Scatter plot of the RMT asymmetry index against the affected-side StartReact effect (VART – VSRT) showing no correlation. **(B)** StartReact effect on the unaffected and affected sides of patients; grey lines connect the two sides of each participant. **(C)** StartReact effect in healthy controls (right hand) and in patients (affected side). **(D)** StartReact effect in healthy controls (right hand) and in patients with more severe upper-limb impairment (affected side; FMA-UE motor score at or below the cohort median of 35). Boxes show the median and interquartile range, whiskers the range excluding outliers, and points individual participants.

The StartReact effect did not differ between the affected and unaffected sides (mean difference, affected − unaffected, 2.6 ms, 95% CI −8.1 to 13.3; median difference 2.4 ms; paired t (54) = 0.48, p = 0.631; N = 55 pairs; Figure 3B).

The affected-side StartReact effect of patients did not differ from that of healthy controls (Mann–Whitney U = 352, z = 0.95, p = 0.342; N = 56 patients and 15 controls; 95% CI of the median difference, patients − controls, −9.8 to 22.8 ms; Figure 3C). However, patients with more severe upper-limb impairment (FMA-UE motor score ≤ 35; N = 23) had a larger affected-side StartReact effect than healthy controls (median 57.5 vs 34.7 ms; Mann–Whitney U = 97, z = 2.24, p = 0.025; 95% CI of the median difference, patients − controls, 2.6 to 36.7 ms; Figure 3D).

In addition, we compared the StartReact effect between the affected and unaffected sides in patients with more severe upper-limb impairment (FMA-UE motor score ≤ 35). In this subgroup also, the StartReact effect did not differ between the two sides (mean difference, affected − unaffected, 13.5 ms, 95% CI −5.0 to 31.9; paired t (20) = 1.53, p = 0.143; N = 21 pairs).

## Discussion

This study examined, in the same group of patients with chronic ischemic stroke, how corticospinal and reticulospinal-associated measures relate to upper-limb motor impairment. Greater RMT asymmetry, reflecting poorer corticospinal function, was associated with greater upper-limb impairment and weaker grip. A larger StartReact effect was also associated with greater impairment and weaker grip. The two measures were not linearly related to each other. The StartReact effect did not differ between the affected and unaffected sides, and overall, it did not differ from that of healthy controls. However, patients with more severe upper-limb impairment had a larger StartReact effect than healthy controls, which was similar on both sides. These results suggest that upper-limb impairment after stroke relates to both CST damage and RST upregulation, and the extent of RST upregulation may not depend on CST damage alone. Together, these findings suggest that the CST and the RST relate to upper-limb impairment in different ways after stroke.

### Corticospinal function and upper-limb impairment

The association between RMT asymmetry and upper-limb impairment agrees with a large body of evidence linking CST integrity to motor outcome after stroke. Imaging studies have shown that greater CST damage is associated with greater impairment (3-5), and preserved corticomotor responses identify patients with good recovery potential (6, 34-36). Our study supports this relationship using a continuous asymmetry index rather than simply classifying patients by the presence or absence of MEPs. Because motor threshold varies between people for reasons unrelated to stroke, such as scalp-to-cortex distance (37), comparing the affected with the unaffected side provides a within-person measure that is less influenced by such differences. RMT asymmetry was associated not only with the overall motor score but also with grip strength. This suggests that residual corticospinal output contributes to force production as well as to the fine, selective control for which the CST is best known (8, 13, 38).

### Reticulospinal and corticospinal contributions to upper-limb impairment

A larger StartReact effect was associated with poorer upper-limb function and weaker grip. This replicates our earlier findings in chronic stroke (27, 28) and agrees with reports that startle-related effects on the paretic arm are more prominent in more impaired individuals (7, 39, 40).

One possible explanation involves cortical input to the RST. Reticulospinal activity is strongly shaped by corticoreticular projections from the premotor, supplementary and primary motor areas (41, 42), and in monkeys, widespread corticoreticular fibers from M1 and SMA bilaterally converge on the reticular formation (43), with the contributing cortical regions extending beyond these two (41). After stroke, these supramedullary projections may be damaged or may rely more on the contralesional hemisphere (17, 44, 45). The StartReact effect may therefore reflect the combined state of the cortex and brainstem, rather than brainstem output alone. This is supported by evidence that disrupting the motor cortex with TMS delays startle-triggered movements (46, 47).

In our earlier work, we proposed that more severely impaired patients have greater CST damage, which leads to compensatory upregulation of the RST (27, 28), implying a linear relationship between the two. This could not be tested then because CST integrity was not estimated objectively with TMS. In the present study we tested both directly. RMT asymmetry was not linearly correlated with the StartReact effect, although both were related to upper-limb impairment. In light of this result, our updated hypothesis is that RST upregulation is more closely linked to damage to the widespread supramedullary fibres connecting the cortex with the reticular formation than to damage of the CST and its collaterals specifically. More severely impaired patients are likely to have greater damage to these corticoreticular connections, which may lead to greater upregulation of RST output. The degree of upregulation may not depend on CST damage alone, because other fibres also modulate RST output (41). We were not able to measure the overall (cumulative) integrity of the corticoreticular connections; this hypothesis could possibly be tested in future work with diffusion tractography.

The lack of a linear relationship is also in keeping with the two pathways contributing to different aspects of motor control. The CST appears particularly important for precise finger movements, and the RST for strength and whole-hand grasp (13, 14, 48). The two pathways also make distinct contributions to arm and hand muscles after spinal cord injury (29, 30). In chronic stroke, stronger CST projections are linked to better motor control and stronger corticoreticulospinal projections to greater strength (20).

In this cross-sectional design, the negative association between RST activity and upper-limb function also cannot exclude the possibility that higher RST activity is itself detrimental to upper-limb function, for example through greater spasticity, stronger flexor synergies or loss of finger individuation. A non-linear relationship between CST damage and RST upregulation also cannot be excluded. Reticulospinal excitability has been reported to increase mainly after severe CST damage (7), and a threshold effect may have been missed because patients without a measurable affected-side motor threshold could not be included. It is unlikely that the larger StartReact effect – the difference between visual-auditory and visual-startle reaction times - simply reflects slower voluntary reactions in patients with more CST damage, because the StartReact effect did not increase with RMT asymmetry.

### StartReact effect on patients and healthy controls

Overall, the affected-side StartReact effect of patients did not differ from that of healthy controls. This contrasts with our earlier finding of a larger StartReact effect after stroke (27). One explanation is that our inclusion criteria required active wrist and finger extension, so that the patients in this study were relatively mildly impaired. As noted above, reticulospinal enhancement appears to be most evident after severe CST damage. In line with this, patients with more severe upper-limb impairment had a StartReact effect about 23 ms larger than that of healthy controls. This finding agrees with the inverse association between the StartReact effect and FMA-UE score in the whole cohort, with greater reliance on reticulospinal pathways in more impaired patients (7, 27). Reticulospinal enhancement after stroke therefore appears not to be universal but to depend on the severity of motor impairment.

The enhanced StartReact effect was also similar on the affected and unaffected sides, both in the whole cohort and in patients with more severe impairment. This is consistent with the bilateral organization of the reticulospinal system. Reticulospinal neurons project to both sides of the spinal cord and thereby influence arm muscles bilaterally (9, 49), and the reticular formation receives input from both hemispheres (43). In line with this, stretch reflex excitability is increased in both the paretic and non-paretic arms of stroke survivors (50). Where reticulospinal output is enhanced, the enhancement may therefore involve both sides.

### Clinical implications

Assessing both descending systems may give a fuller picture of residual impairment than either measure alone. Corticospinal measures already help to predict recovery potential (34, 35). The StartReact effect, which requires only simple equipment, may add additional information about reliance on reticulospinal pathways. Because a greater StartReact response was associated with poorer function and StratReact was enhanced mainly in more severely impaired patients, strategies that simply increase reticulospinal drive may not be appropriate for all patients. Therapies that strengthen residual corticospinal output may suit patients with relatively preserved CST function, whereas approaches that engage reticulospinal pathways in a targeted way may be more relevant for those with severe CST damage (51, 52). These possibilities need to be tested in prospective studies.

### Strengths and limitations

The strengths of this study include a relatively large sample, assessment of both pathways in the same individuals, comparison with age-matched healthy controls, and correction for multiple comparisons.

A limitation of this study is that RMT asymmetry required a measurable motor threshold on the affected side. Patients with the most severe CST damage, who are the group most likely to rely on the RST, were therefore under-represented, which may have weakened the relationship between the two measures.

## Conclusion

In chronic ischemic stroke, poorer corticospinal function and greater reticulospinal involvement were each associated with greater upper-limb motor impairment and weaker grip but were not associated with each other. This suggests that the two pathways contribute independently to post-stroke motor control. Although the StartReact effect of patients overall did not differ from that of healthy controls, patients with more severe upper-limb impairment showed a larger response, which was similar on the affected and unaffected sides. This suggests that reticulospinal enhancement after stroke depends on the severity of impairment and may involve both sides. Combined assessment of corticospinal and reticulospinal-associated measures may therefore offer a more complete, objective characterization of upper-limb impairment after stroke and could help guide pathway-targeted rehabilitation in future.

## Data Availability

The data are not publicly available because of participant confidentiality; de-identified data may be made available upon reasonable request to the corresponding author following peer-reviewed publication, subject to approval by the Institutional Ethics Committee of Institute of Neurosciences Kolkata.

## Acknowledgement

The authors sincerely thank all the participants and their caregivers for their time, cooperation, and participation in the study. The authors acknowledge the institutional and research support provided as part of the Indian Council of Medical Research (ICMR)-funded small grant program.

